# Safety and immunogenicity of a vaccine against coxsackieviruses B (PRV-101) – follow-up of the first-in-human phase 1 trial

**DOI:** 10.64898/2026.02.05.26345529

**Authors:** Jutta E. Laiho, Jussi P. Lehtonen, Leena Puustinen, Susanna Kääriäinen, Taina Härkönen, Sami Oikarinen, Francisco León, Miguel Sanjuan, Mika Scheinin, Mikael Knip, Heikki Hyöty

## Abstract

**Background:** Coxsackie B viruses cause acute infections and have been linked to chronic diseases like cardiomyopathies, type 1 diabetes, and celiac disease. Despite their clinical significance, no vaccines exist for coxsackie B virus types. PRV-101, a new candidate vaccine covering five coxsackie B virus types, showed good immunogenicity and tolerability in a phase 1 trial (PROVENT).

**Methods:** We conducted an extended follow-up of the PROVENT trial to assess the long-term immune response and safety of PRV-101. A total of 26 participants from the original cohort (n=32) were enrolled for additional testing approximately two years post-immunization (11 high-dose, 10 low-dose, and 5 placebo). Coxsackie B virus -specific antibody responses were measured and compared to earlier time points.

**Results:** PRV-101 was safe with no late adverse effects or emergence of autoantibodies linked to type 1 diabetes or celiac disease. Neutralizing virus antibodies remained elevated, with a clear dose-dependent response. In the high-dose group, antibodies against all coxsackie B virus types reached presumably protective levels, except for coxsackie B virus 2, where two participants turned seronegative. ELISA tests confirmed elevated antibody levels against coxsackie B virus proteins.

**Discussion:** These results suggest that PRV-101 induces durable antibody responses lasting at least two years. The findings support the continued development of PRV-101 for preventing both acute coxsackie B virus infections and chronic diseases like type 1 diabetes and celiac disease.

## 1. BACKGROUND

Coxsackievirus B (CVB) infections are common in all age groups. They can cause a variety of symptoms and conditions, ranging from mild common cold -type respiratory disease to severe illnesses, such as meningitis, encephalitis, myocarditis, and hand, foot and mouth disease (1). In young infants, severe CVB infections may sometimes be fatal. In addition, CVBs have been linked to chronic diseases such as chronic cardiomyopathies, type 1 diabetes and celiac disease, where persistent CVB infection in the heart, pancreas and gut mucosa, respectively, may be involved (2,3). There are more than 110 human-infecting enterovirus types that are divided into four species, *Enterovirus A-D*. CVBs belong to *Enterovirus B* and include six types. The medical significance of these infections is demonstrated by the fact that among the >110 different enterovirus types CVBs have continuously been among those 15 virus types that have most frequently led to health care contacts in the US (4,5). In spite of the significant disease burden, no vaccine is available for the prevention of CVB-related diseases.

We have started a clinical development program aimed at a pentavalent human CVB vaccine capable of protecting against CVB-associated diseases. This formalin-inactivated whole-virus vaccine (PRV-101) includes the most frequent CVB types (CVB1-5) and contains no adjuvant. PRV-101 was recently tested in a first-in-human phase 1 trial (PROVENT). Three immunizations one month apart did not lead to vaccine-associated severe adverse effects (6). Three doses of PRV-101 induced robust dose-dependent antibody responses against each of the five CVB types included in the vaccine. The last study visit occurred 24 weeks after the last immunization. At that time, the vaccine-induced antibody levels already showed some decline, still remaining at high levels, exceeding those considered protective against enterovirus infections. The aim of the present study was to evaluate the longer-term safety and immune response of PRV-101 by analyzing the antibody levels at a follow-up visit, approximately two years after the last immunization. Here we report that PRV-101-induced neutralizing antibody responses persisted throughout this longer observation period at levels which are considered protective against CVB infections.

## 2. SUBJECTS AND METHODS

### Study design and trial participants

This is an Investigator-initiated follow-up study (IIS) of safety and long-term immune response in participants of the PROVENT trial, a first-in-human phase 1 trial of PRV-101, a CVB vaccine candidate (hereafter termed PROVENT-IIS), EudraCT number 2022-004174-39, funded by Provention Bio (later acquired by Sanofi). Principal investigator was professor Heikki Hyöty (Tampere University, Finland) and Tampere University, Tampere, Finland was the study sponsor. Recruitment of the study subjects was carried out by Clinical Research Services Turku (CRST Oy) (Turku, Finland). Ethical approval was obtained from the National Committee on Medical Research Ethics (TUKIJA), and the trial protocol was approved by the Finnish Medicines Agency (Fimea).

The recruitment and randomization of the 32 participants of the original PROVENT trial has been described previously (6). Briefly, participants were randomized into three parallel dosing cohorts as follows: low dose (100 µl) PRV-101 (n=12), high dose (500 µl) PRV-101 (n=12), placebo (n=8). In the present study, we invited all 32 participants to attend an additional study visit (IIS visit) by contacting them by phone. The IIS visit occurred 69-88 weeks after the end of study (EOS) visit which corresponds to 101-120 weeks (mean 108 weeks = 27 months) after the first PRV-101 dose and 93-112 weeks (mean 100 weeks = 23 months) after the last PRV-101 dose. The PROVENT trial profile amended with PROVENT-IIS is shown in Fig 1.

**Fig. 1.**
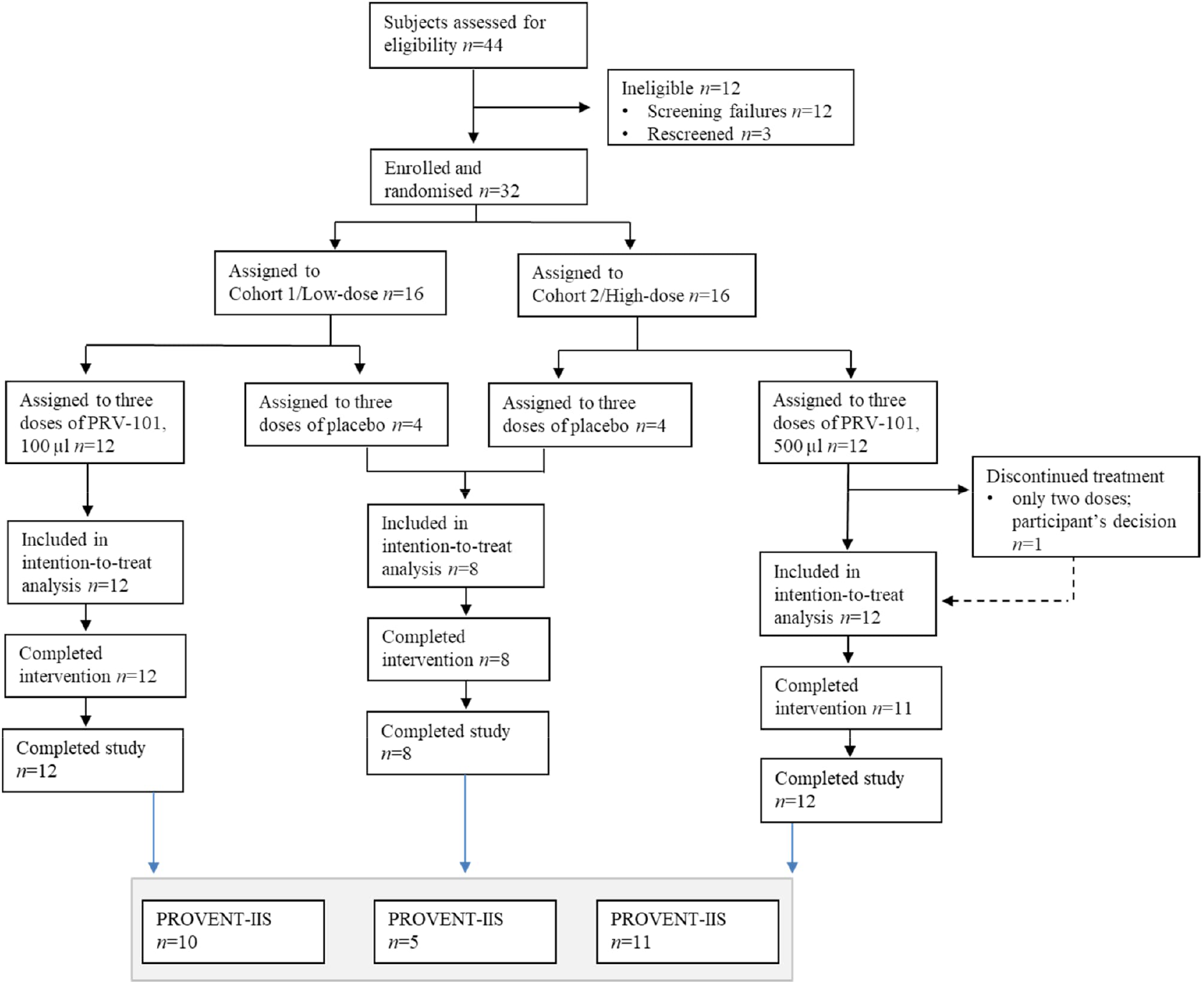
PROVENT trial profile, and participants enrolled in PROVENT-IIS from the three trial arms.

Altogether 26 subjects consented to PROVENT-IIS and were invited to visit CRST’s clinic. The subjects belonged to all three trial arms representing placebo (n=5), low dose (n=10) and high dose (n=11) of PRV-101. At CRST, the health status of the participants was recorded by a questionnaire and a blood sample was drawn. The serum fraction was used for the analysis of CVB antibodies and for autoantibodies associated with type 1 diabetes or celiac disease. Fig. 2 presents a schematic illustration of the study timeline and provides a summary of the age and sex distribution of the study participants.

**Figure 2.**
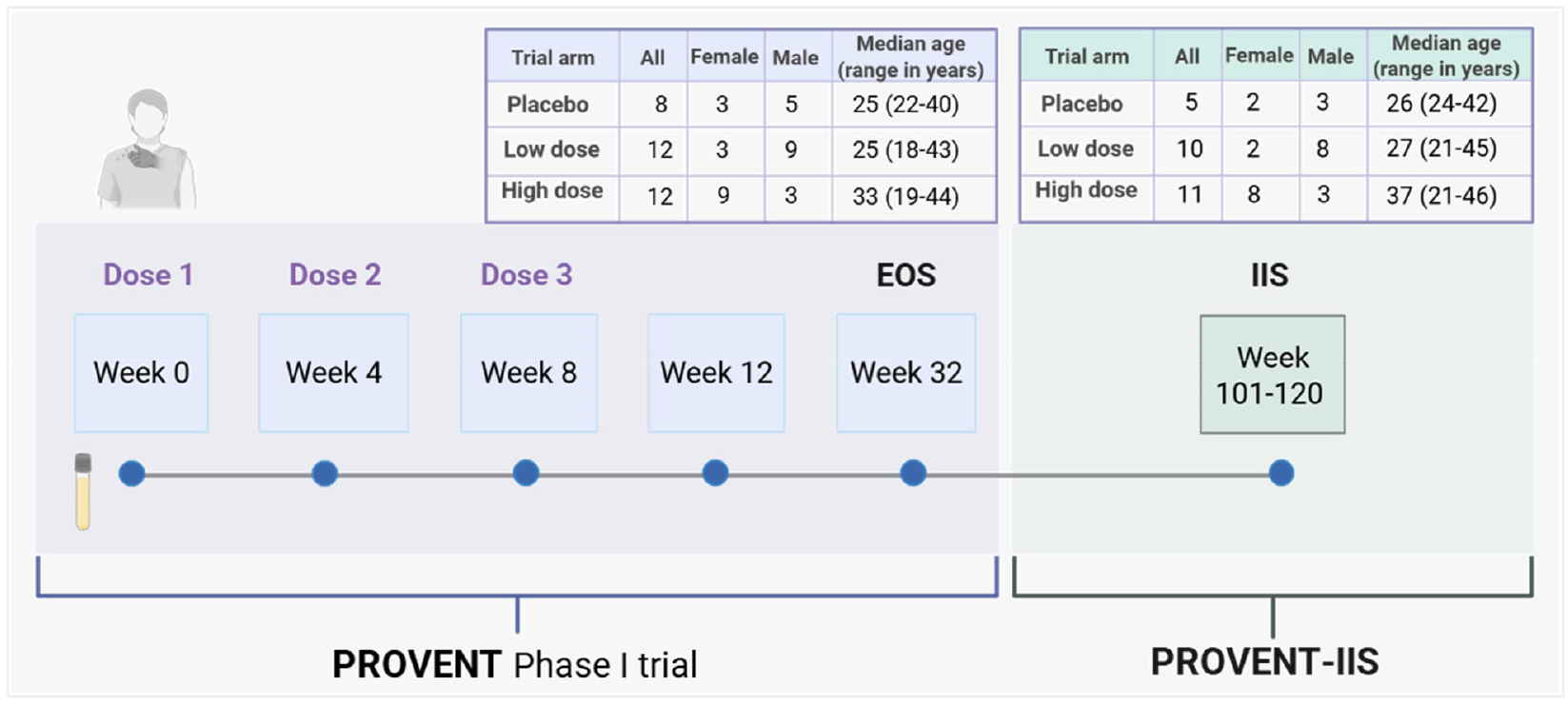
Schematic illustration of the PROVENT trial and PROVENT-IIS designs. In the original PROVENT Phase I trial, participants were given three i.m. injections of PRV-101 or placebo at intervals of 4 weeks and followed afterwards for safety for 24 weeks until the end-of-study (EOS) visit. Blood and nasal swab samples were also collected at each visit. We were able to recruit 26 of the PROVENT participants to the PROVENT-IIS. An additional blood sample was collected at the IIS time point, 101 to 120 weeks after the first injection of PRV-101 or placebo. For the PROVENT trial, virus antibody analyses and type 1 diabetes and celiac disease autoantibody analyses were carried out from week 0, 4, 8, 12 and EOS samples and, for PROVENT-IIS, from the EOS and IIS samples. Age of the participants in both studies is calculated at the time of consent for the PROVENT Phase I trial.

### Virus antibodies

We carried out virus antibody analyses from the collected IIS samples of the 26 participants as well as from their corresponding EOS samples, collected in the main PROVENT trial. The analyses were done in the same virus laboratory at Tampere University, using the same assays as in the main PROVENT trial (6). Briefly, virus type-specific antibodies against each of the CVB1-5 strains included in PRV-101 were analyzed using plaque reduction neutralizing antibody assays. The end-point neutralizing antibody titer was determined by 2-step dilution series of serum samples.

The ELISA IgG and IgM class CVB antibody levels in serum were analyzed using the same commercial antibody assays as in the main PROVENT trial [SERION ELISA classic Coxsackievirus IgG (ESR134G), IgM (ESR134M) Serion GmbH, Würzburg, Germany] according to the manufacturer’s instructions with a single serum dilution (IgM 1:100, IgG 1:500). In these tests, the antigen is a mixture of recombinant antigens derived from conserved and subtype specific epitopes of the VP1 proteins of coxsackieviruses B1, B3 and B5. Antibody units were calculated according to the manufacturer’s instructions.

### Autoantibodies

We analyzed the autoantibodies against type 1 diabetes associated islet cell antigens [antibodies to insulin (IAA), truncated GAD65 (GADA), islet antigen 2 (IA-2A), and zinc transporter 8 (ZnT8A)] in the PEDIA laboratory, University of Helsinki and celiac-disease associated anti-transglutaminase IgG and IgA class antibodies in Turku University Hospital, similarly to the procedures in the main PROVENT trial (6,7).

### Statistical analyses

Virus neutralizing antibodies and autoantibodies were analyzed both from the IIS time point as well as from the EOS sample of the main PROVENT trial. This enabled the adjustment of the virus antibody levels detected in the samples collected in the current study (weeks 101-120 after baseline) to the levels detected in the original analyses of the EOS samples. The adjustment, referred to as ‘EOS-adjusted’ hereafter, was done using the following formula:

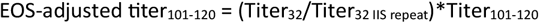

where the numbers indicate the weeks after the first dose of vaccine, EOS time point being week 32 and IIS time point week 101-120. These adjusted antibody levels were used when overall kinetics of antibody levels were analyzed from the baseline sample and all following samples, including the IIS follow-up sample.

Data are presented using descriptive statistics. Antibody levels were log-transformed to present the variation in the vaccine-induced antibody responses; log2-transformed for virus neutralizing titers, and ln-transformed for ELISA antibodies. Mean and median values of the log-transformed antibody levels were used in representative figures. R version 4.2.1 (www.r-project.org, accessed 23 June 2023) was used for statistical analysis.

## 3. RESULTS

### Health status of trial participants

The health status of all trial participants was evaluated by a questionnaire at CRST during the IIS study visit. None of them had developed type 1 diabetes, celiac disease or any other chronic disease after the EOS visit of the main PROVENT trial. No new chronic medications had been started, either.

### Autoantibodies

All IIS participants remained negative for all tested type 1 diabetes -related islet autoantibodies (IAA, GADA, IA-2A, ZnT8A) and celiac disease -related IgG and IgA class anti-transglutaminase autoantibodies.

### Neutralizing antibodies against different coxsackievirus B types

Analysis of CVB neutralizing antibody titers over the whole trial period showed that PRV-101 -induced neutralizing antibody responses persisted at the IIS time point (Fig. 3, Supplementary Fig. 1, Supplementary Table 1), remaining at elevated levels also in participants who were baseline seronegative (titer <4) for the tested CVB type (Fig. 4, Supplementary Fig. 2, Supplementary Table 2), including also those five participants who were baseline seronegative for all five CVB types (Supplementary Fig. 3).

**Figure 3.**
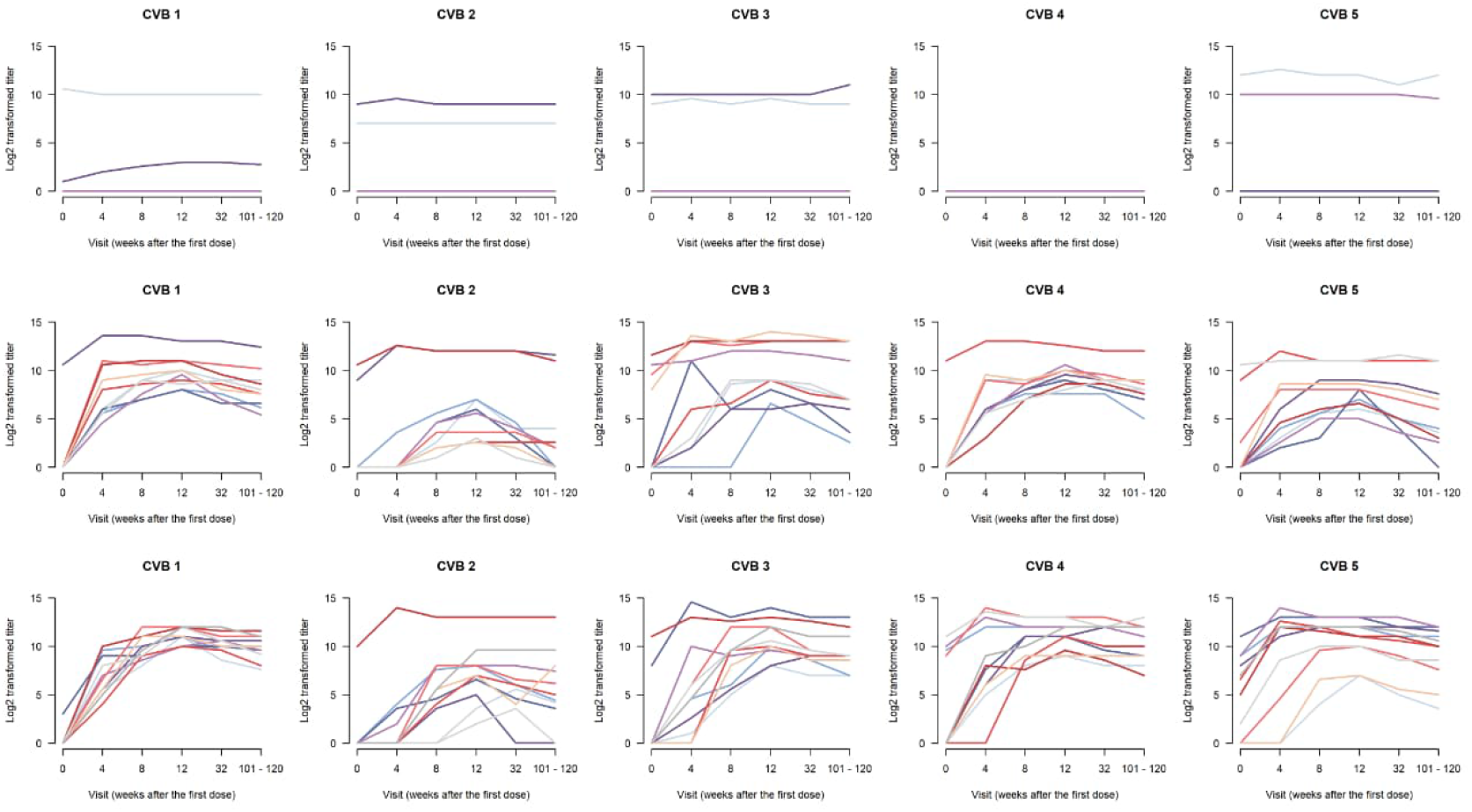
Neutralizing antibody titers (log scale) against CVB1-5 viruses in individual PROVENT-IIS trial participants. Each line represents one individual, its respective color staying the same through the virus antibody panels. The top panel represents the placebo group (n=5), the middle panel the low dose PRV-101 group (n=10) and the bottom panel the high dose group (n=11). The IIS visit occurred 101-120 weeks after the first PRV-101 dose. The presumably protective titer 8 equals to log2 value 3. All participants of the placebo group who were seronegative at baseline remained seronegative throughout the follow-up.

**Figure 4.**
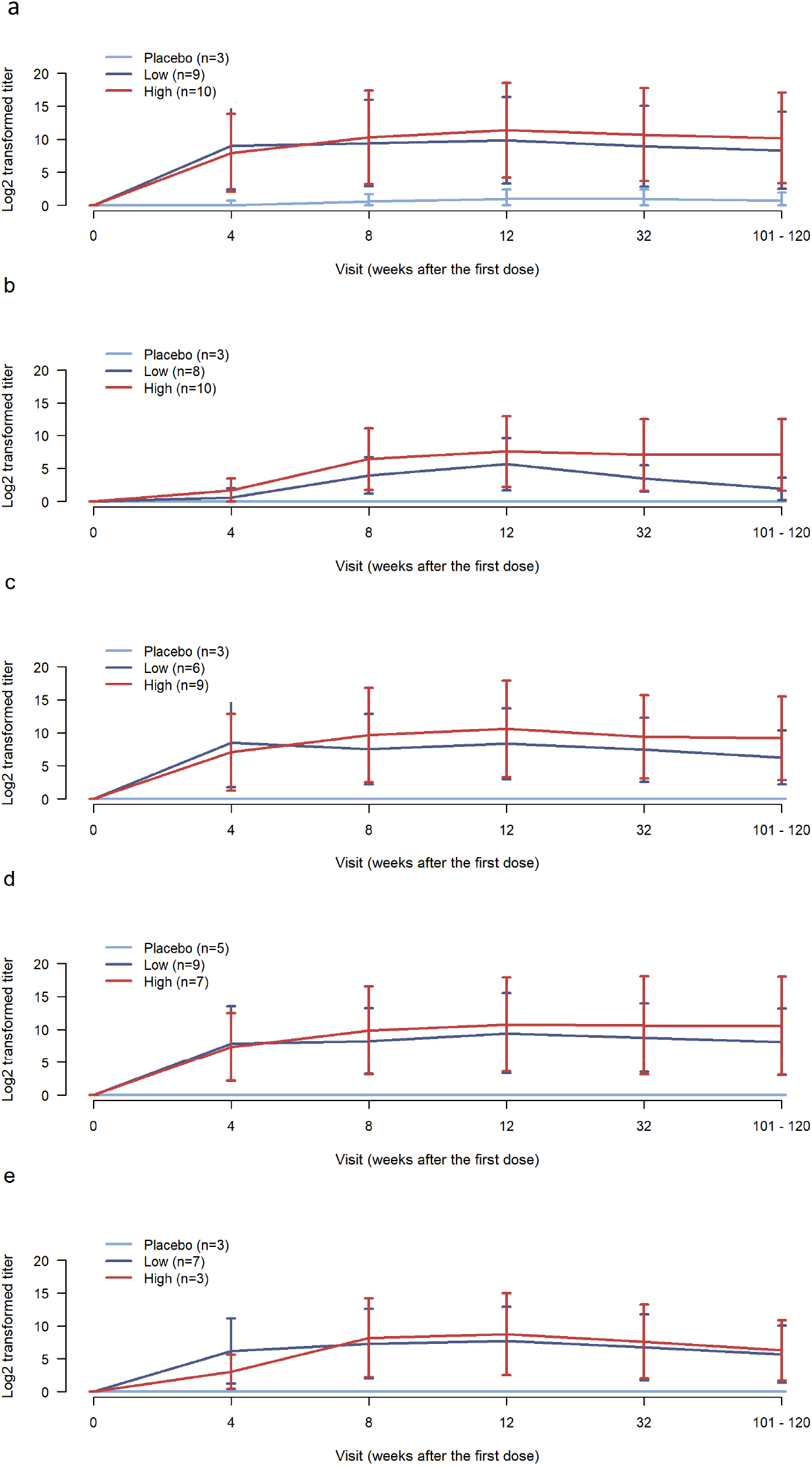
Neutralizing antibody titers (mean +/-SD in logarithmic scale) against CVB1-5 viruses in PROVENT-IIS trial participants who were initially seronegative at baseline for each tested CVB type (numbers of subjects are shown in each panel). a) CVB1; b) CVB2; c) CVB3; d) CVB4; e) CVB5. The IIS visit took place 101-120 weeks after the first PRV-101 dose.

### Levels of the neutralizing antibody titers in the high and low dose groups

Among the baseline seronegative participants, clear dose-dependency of the antibody response was observed. The higher dose of PRV-101 led to higher antibody levels at the IIS timepoint when compared to antibody levels in the lower dose group (Fig. 4). This difference was most obvious for CVB2 which induced generally smaller antibody responses compared to the other CVB components of PRV-101 (Supplementary Table 1, and Supplementary Fig. 3 and 4). All baseline seronegative participants in the placebo group remained seronegative throughout the follow-up.

In the high-dose group, all baseline-seronegative participants had IIS antibody titers of 4 or higher for CVB1, CVB3, CVB4 and CVB5, while CVB2 titers had declined below this level in two of the ten baseline CVB2 seronegative subjects (Fig. 3, Supplementary Fig. 3). IIS titers remained at ≥ 4 for all CVB types in both of the two high-dose participants who were baseline seronegative for all five CVBs (Supplementary Fig. 3 and 4).

In the low-dose group, all baseline seronegative participants had IIS antibody titers ≥4 for CVB1, CVB3 and CVB4, while CVB2 titers had decreased below 4 in 4 of the 8 such participants, and CVB5 antibodies had decreased below 4 in 1 of the 7 such participants. All three low-dose participants who were baseline seronegative for all five CVB types had IIS titers of ≥ 4 against CVB1, CVB3, and CVB4, while this was the case for 1 of participants for 3 CVB2 and 2 of 3 participants for CVB5 (see Supplementary Fig. 3 and 4).

Since neutralizing antibody titers of ≥ 8 have been linked to protection against enterovirus infections (e.g. regarding polioviruses) (8), we analyzed the proportion of baseline seronegative participants whose titers remained at this level still at the IIS time point. In the high-dose group, the results were identical with those for titers ≥ 4, whereas in the low-dose group, all baseline seronegative participants had IIS antibody titers ≥ 8 for CVB1 and CVB4, while CVB2 titers had decreased below 8 in 7 of 8 such individuals, CVB3 titers in 1 of 6 and CVB5 antibodies in 2 of 7 such individuals. In addition, the three low-dose participants seronegative at baseline for all five CVB types had IIS titers ≥ 8 against CVB1 and CVB4 while this was true for 1 of 3 such individuals for CVB2, 2 of 3 for CVB3, and 2 of 3 for CVB5 (see Supplementary Fig. 3 and 4).

### Coxsackievirus B antibodies in the ELISA assay

In ELISA assays, the levels of IgG class CVB antibodies decreased gradually after the 12 weeks’ timepoint (i.e. one month after third vaccine dose) both in the high and low PRV-101 dose groups but still remained clearly above baseline levels at the IIS time point (Fig. 5, Supplementary Table 1 and 2, Supplementary Fig. 5 and 6). In contrast, IgM antibodies decreased more steeply, declining near baseline levels at the IIS time point. In the placebo group, no clear changes occurred in IgG or IgM antibody levels.

**Figure 5.**
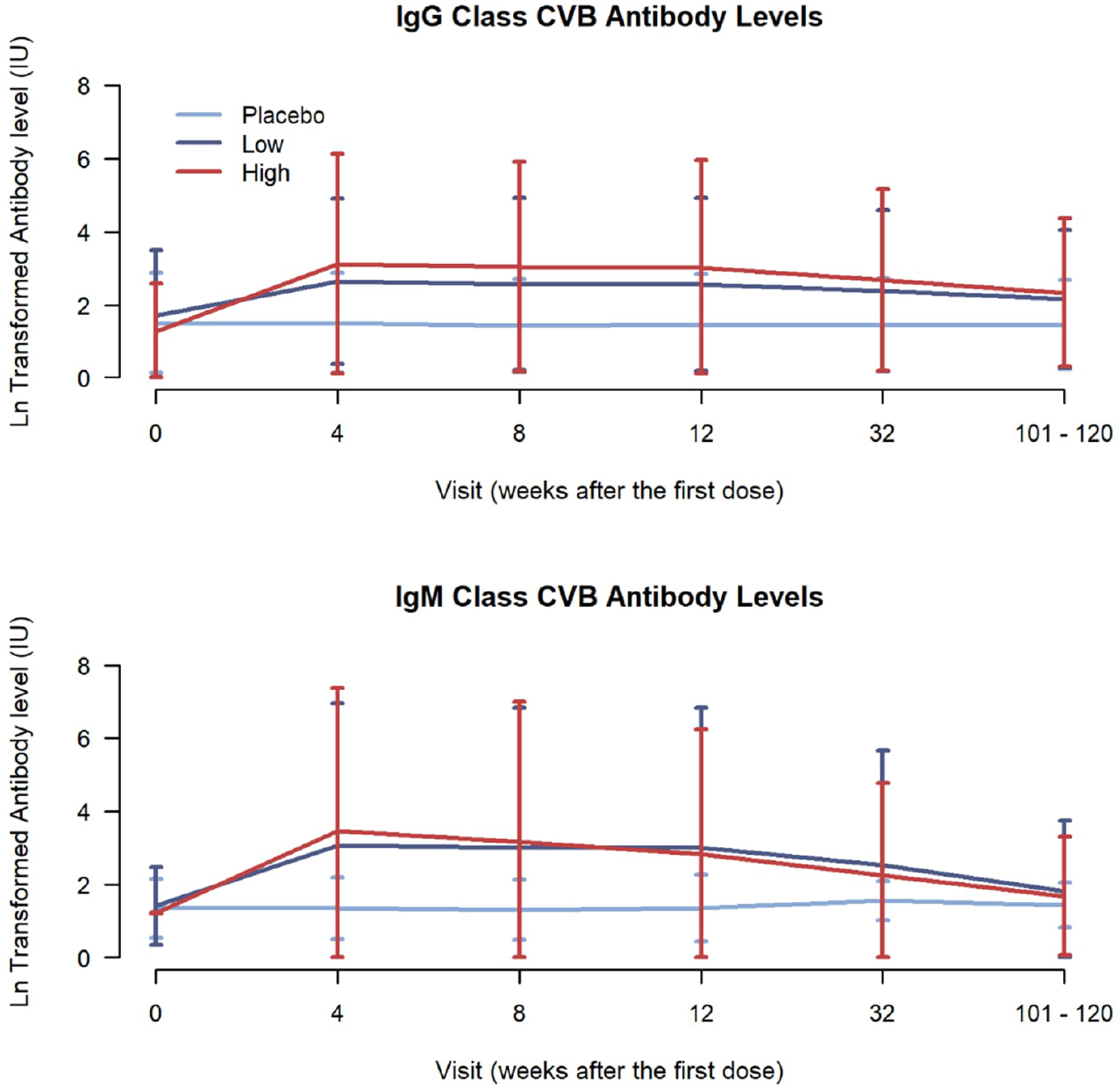
The levels of IgG and IgM antibodies against CVB antigen in PROVENT-IIS trial participants, using Serion IgG and IgM ELISA assays.

## 4. DISCUSSION

The results of the present follow-up study indicate that PRV-101 can induce long-lasting antibody responses in initially CVB seronegative individuals. Neutralizing CVB antibodies that correlate with protection against CVB infections remained elevated until approximately two years after the last dose of PRV-101. Clear dose-dependency was also seen – the participants of the high dose group had higher neutralizing antibody levels, and they were more frequently seropositive (titer ≥ 4) at this late time point when compared to the participants of the low dose group. In addition, in the high dose group all baseline seronegative participants reached presumably protective titers of ≥ 8 against CVB1, CVB3, CVB4 and CVB5, and 80% of them also for CVB2. Titers of ≥8 have been considered protective in previous poliovirus studies (8). Similar dose-dependent patterns were also seen in IgG class CVB antibodies when measured with ELISA. Overall, these results suggest that three immunizations with the higher tested dose of PRV-101 lead to robust and sustained immune responses, including the induction of neutralizing antibodies that mediate protection against CVB infections. The observation that PRV-101-induced antibodies remain elevated over such a long time suggests efficient induction of memory B-cells by PRV-101 (9).

The results are well in line with the previous experience from other inactivated enterovirus vaccines. The widely used inactivated poliovirus vaccine (IPV) generates efficient protection against polio paralysis that lasts for decades (10). In addition, several clinical trials and post-licensing studies have shown that similar formalin inactivated vaccines against enterovirus 71 induce robust and long-lasting neutralizing antibody responses providing good protection against enterovirus 71 infections (11). Thus, it seems that formalin inactivated whole-virus vaccines work well in immunization against several types of enteroviruses.

The antibody responses persisted also in the lower dose group, but the response to CVB2, towards which the response was initially weaker than the responses against the other CVBs, waned in 7 of the 8 individuals who were CVB2 seronegative at baseline. Some individual variation in the waning of titers below titer 8 was also seen for CVB3 and CVB5 (in 1 of 6 individuals for CVB3 and in 2 of 7 individuals for CVB5, who were seronegative at baseline). As discussed in our previous paper (6), the lower concentration of CVB2 in the PRV-101 vaccine, compared to the other types, is the most likely explanation for the overall smaller responses to CVB2. In any case, the higher dose of PRV-101 seems to be a more favorable option for future clinical trials.

The finding that CVB antibodies remained elevated in immunized subjects who had been CVB seronegative at baseline, including those who had been seronegative for all CVBs, suggests that three immunizations with an appropriate dose of PRV-101 can induce long-term immunity in initially CVB-naïve individuals. However, even though neutralizing antibodies are considered as reliable markers of past CVB infections, it is possible that some of these initially seronegative individuals had been previously exposed to CVBs and had preexisting CVB-specific memory T-cells that can boost PRV-101-induced responses. Therefore, it would be important to carry out a similar trial among young children to confirm the high immunogenicity of PRV-101 in previously unexposed individuals. Young children are also the main target population of PRV-101 since it aims to prevent especially early CVB infections which most frequently cause severe diseases and can also contribute to the development of type 1 diabetes. The interval between the three immunizations was only 4 weeks, which has probably been too short for optimally efficient immunization. Therefore, further studies are needed to test whether longer intervals could lead to stronger immune responses even with the lower dose level of the vaccine. Such studies could also help to address the question of whether the response to CVB2 could be enhanced by modifying the dose or immunization protocol.

PRV-101 is manufactured using similar technology as used in the manufacture of IPVs (highly purified formalin-inactivated viruses). In addition, neither of these vaccines contains adjuvants. Vaccines against another enterovirus, enterovirus 71 (EV-71), have also been widely tested in phase 3 trials showing excellent efficacy and safety, and one of these vaccines is licensed in China (11,12). These monovalent EV-71 vaccines have also been manufactured using similar formalin-inactivation technology as PRV-101. However, in contrast to PRV-101, they contain an alum adjuvant. Our preclinical studies in non-human primates showed that alum adjuvant did not augment the sustainability of neutralizing antibody response induced by a formalin-inactivated prototype vaccine containing all six CVB types (13). Based on these facts it seems that enterovirus vaccines that are based on formalin-inactivated whole virus vaccine technology are efficient and safe in humans and can induced robust and sustained immune responses even without any adjuvant.

All IIS participants remained negative for all four type 1 diabetes -associated autoantibodies and celiac disease associated transglutaminase autoantibodies. None of them had developed type 1 diabetes or celiac disease, or any other chronic illness. Thus, there was no indication that immunization with PRV-101 could increase the risk to developing autoimmune conditions. This is an important result for the future development of PRV-101, particularly since almost a half of IIS study participants carried HLA-conferred genetic susceptibility to type 1 diabetes or celiac disease (6). Of note, PRV-101 does not contain any adjuvant or infective viruses. These are also important safety aspects of PRV-101 when considering immunizations among individuals who carry increased genetic risk to develop these or other autoimmune diseases. The safety of PRV-101 is also supported by knowledge that the current inactivated poliovirus vaccine, which is produced using similar formalin-inactivation technology, is considered as one of the safest vaccines ever developed. In any case, the favorable results together with the relatively small number of participants of this phase 1 trial argue for larger trials to get more detailed information about tolerability, safety and immunogenicity of PRV-101.

The Serion ELISA assays used to study immunogenicity of PRV-101 are based on recombinant VP1 proteins of CVB1, CVB3 and CVB5, and are able to detect antibodies which cross-react between CVB serotypes and are therefore not specific for the individual CVB types that are used as antigen. The assays detected PRV-101 induced antibodies well, indicating that they are suitable for the evaluation of the immunogenicity of PRV-101 in human trials. In addition, VP1 protein seems to be an important target for PRV-101 induced antibodies.

In summary, the results of this investigator-initiated study indicate that immunization with PRV-101 was not associated with any late-appearing adverse effects during an observation period that lasted approximately two years after the first administration of PRV-101. In addition, the CVB neutralizing antibody responses that were induced by three immunizations by the higher dose of PRV-101 remained at high levels during this extended observation period, still exceeding the presumed protective levels. Altogether, these findings support continuation of the PRV-101 clinical development program and make PRV-101 an attractive candidate for a vaccine that could prevent CVB-associated diseases and possibly also type 1 diabetes and celiac disease. Our next step is to confirm the safety and immunogenicity of PRV-101 in a Phase 1b trial in young children who will be the main target group in clinical trials testing the efficacy of PRV-101 in the prevention of type 1 diabetes and celiac disease.

## Supporting information

Supplemental material PROVENT-IIS

## Data Availability

All data produced in the present study are available upon reasonable request to the authors

## Acknowledgements

The authors would like to thank all the participants who agreed to participate in the study. We also wish to thank the personnel of CRST Oy, as well as the virus laboratory of Tampere University and autoantibody laboratory of the University of Helsinki, for their skilful work.

## Conflict of interest

At the time of the PROVENT trial, MSa and FL were employed by Provention Bio, Inc, a Sanofi company, which was the sponsor of the PROVENT trial. MK and HH are board members and stock owners of Vactech Oy, a Finnish biotech company that has contributed to the early-stage development of the vaccine and participated in the trial design and laboratory analyses. Vactech has recently acquired the rights of PRV-101 and continues its clinical development. SK and MSc are employees of CRST Oy, a clinical contract research organisation that was contracted by Tampere University to perform the PROVENT-IIS study. The authors declare that there are no other relationships or activities that might bias, or be perceived to bias, their work.

## Funding

Provention Bio, Inc., a Sanofi company, funded the original PROVENT study and provided part of the funding of the current study as an unrestricted grant to Tampere University.

## Contribution statement

This investigator-initiated study was based on the initial idea of HH, who was also the Principal Investigator of the study and is affiliated to Tampere University (the sponsor). MK, MSc and HH were responsible for the overall design of the study and FL and MSa for the design of the PROVENT trial. SK and MSc were responsible for the recruitment of study participants and management of the study at CRST Oy. JEL and HH were responsible for project administration in the Tampere virus laboratory and TH and MK in the Helsinki islet autoantibody laboratory. TH was responsible for the autoantibody data analyses, JEL was responsible for the virus-neutralising antibody analyses, and LP for the virus PCR and ELISA analyses. JPL was responsible for the data analyses. JEL and HH were primary contributors in the preparation of the manuscript. All authors reviewed and edited the manuscript and approved the final version. HH is the guarantor of this work and accepts full responsibility for the work and/or the conduct of the study, had access to the data and controlled the decision to publish. Provention Bio helped to fund the study with an unrestricted grant to Tampere University, but did not impose restrictions regarding the submission of the report for publication.

## Data availability

The datasets are available from the corresponding author upon reasonable request

## Alt text

**Fig. 1**. A graph showing the PROVENT trial profile and number participants enrolled in PROVENT-IIS from the three trial arms (High dose, Lod dose and placebo).

**Fig.2**. Schematic illustration of the PROVENT trial and the PROVENT-IIS trial design, showing basic demographics of the participants per trial arm in both PROVENT and PROVENT-IIS, and the intervals of vaccine doses and sample collection points.

**Fig. 3**. Charts showing neutralizing antibody titers against coxsackievirus 1-5 in individual PROVENT-IIS trial participants, starting from the first vaccine or placebo dose (week 0) until the IIS timepoint (week 101-120). Charts are shown per trial arm, and individual lines represent individual participants.

**Fig. 4**. Graph showing neutralizing antibody titers against coxsackievirus 1-5 in PROVENT-IIS trial participants who were initially seronegative at baseline for each tested virus type, starting from the first vaccine or placebo dose (week 0) until the IIS timepoint (week 101-120). The graphs are shown per virus type and each trial arm is presented in one color line (red for High dose; dark blue for Low dose, light blue for placebo).

**Fig. 5**. Graph showing the levels of IgG and IgM antibodies against coxsackievirus B antigens in trial participants. Each trial arm is presented in one color line (red for High dose; dark blue for Low dose, light blue for placebo).

## Notes

### Clinical Trial

EudraCT number 2022-004174-39

### Author Declarations

Ethics Committee of the Hospital District of Southwest Finland gave ethical approval for this PRV-101 vaccine trial work 23 Sep, 2020. This current work is a follow-up of this trial.

