## Supplemental material PROVENT-IIS for "Safety and immunogenicity of a vaccine against coxsackieviruses B (PRV-101) – follow-up of the first-in-human phase 1 trial"

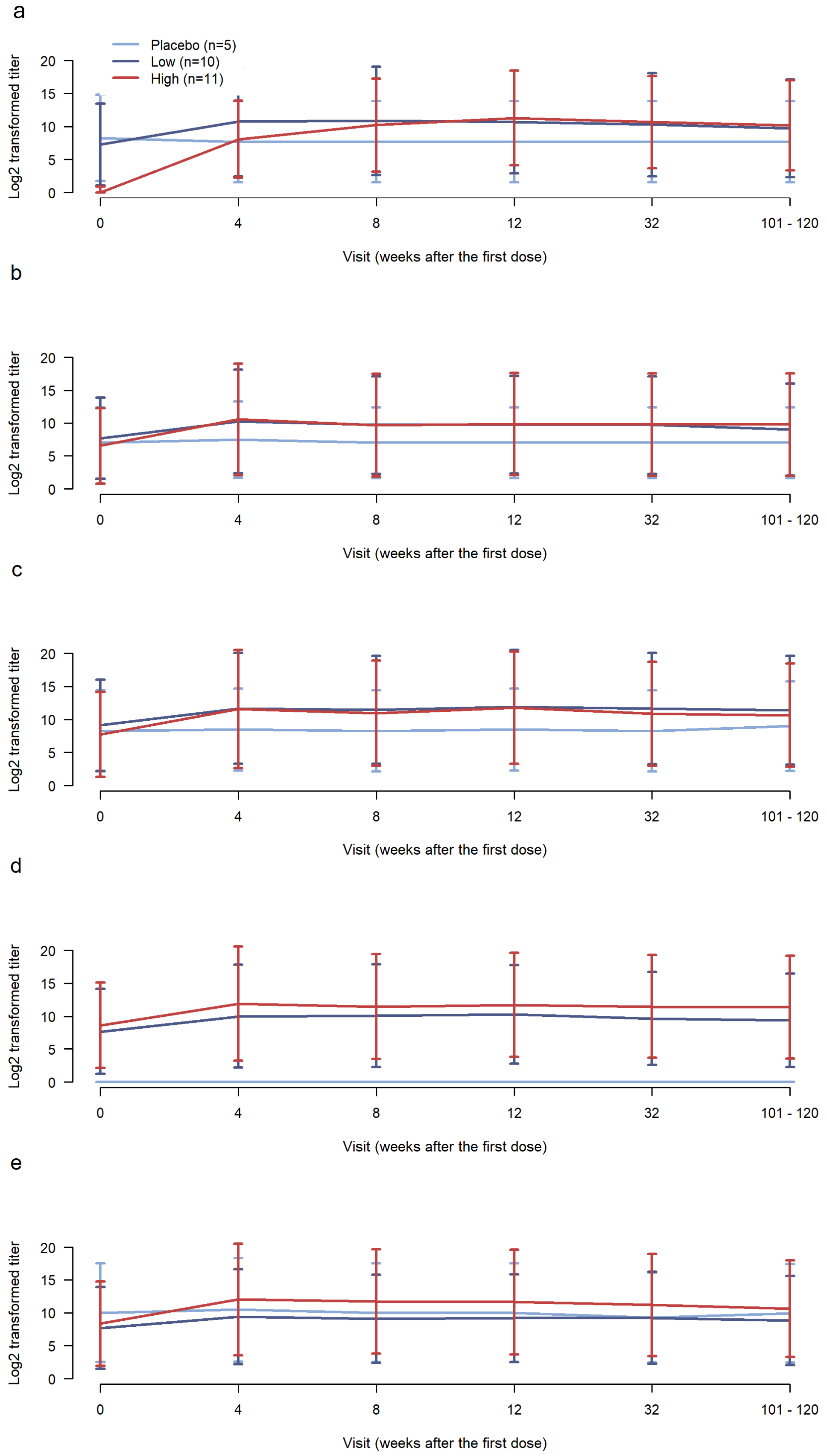


**Supplementary Figure 1.** Neutralizing antibody titers (mean +/- SD in logarithmic scale) against CVB1-5 viruses in the PROVENT-IIS trial participants (n=26) for the tested CVB type (numbers shown in each panel).  a) CVB1; b) CVB2; c) CVB3; d) CVB4; e) CVB5. The IIS visit occurred 101-120 weeks after the first PRV-101 dose

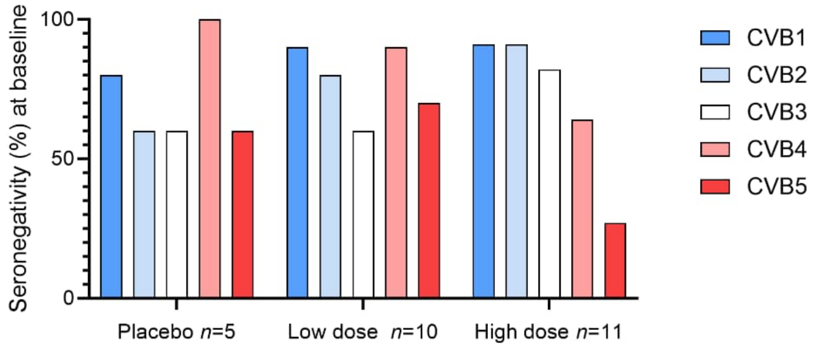


**Supplementary Figure 2.** Proportion of PROVENT-IIS participants assessed as seronegative (titer <4) for neutralizing antibodies at baseline prior to the fist PRV-101 dose against each of the tested CVB types.

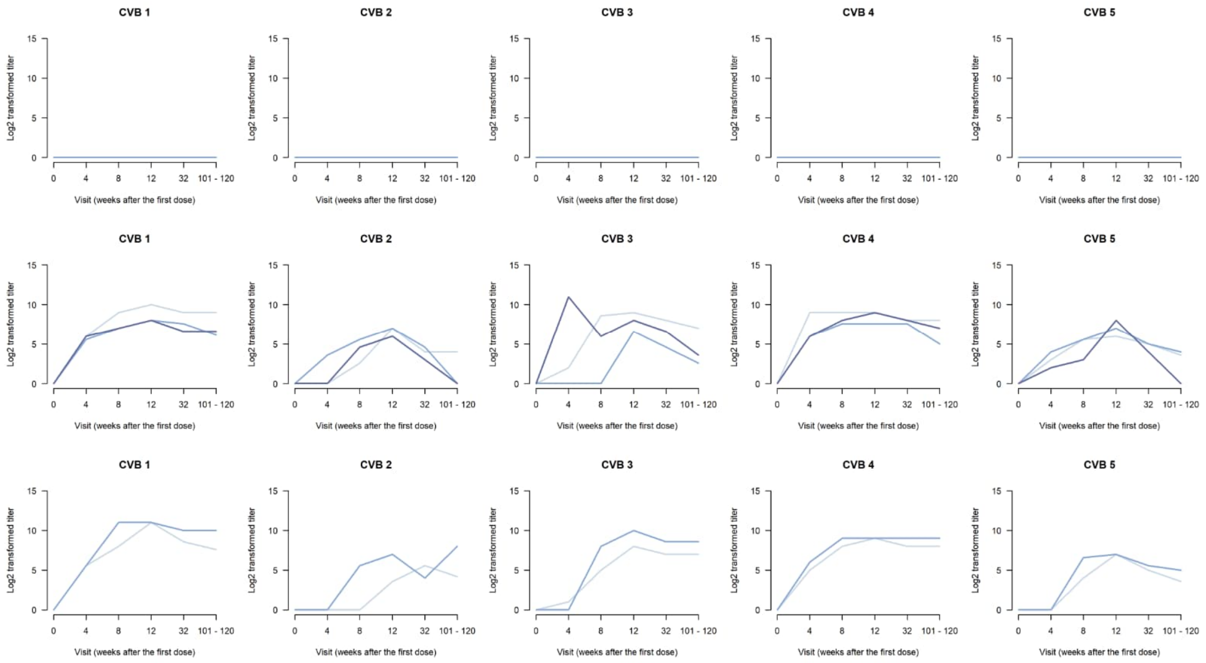


**Supplementary Figure 3.** Neutralizing antibody response to CVB1-5 viruses in those PROVENT-IIS trial participants who tested negative for neutralizing antibodies against all CVB1-5 viruses at baseline. The top panel represents the placebo group (N=2), the middle panel the low dose PRV-101 group (N=3) and the bottom panel the high dose PRV-101 group (N=2). Each participant is presented by an individual line, its respective color staying the same through the virus antibody panels. The presumably protective titer 8 equals to log2 value 3.

**Supplementary Table 1**. Median (range) neutralizing antibody titers (linear scale) against CVB1-5 viruses and median antibody levels against CVB antigens (ELISA) at the PROVENT-IIS time-point. The data has not been adjusted to the EOS time point.

| | | Neutralizing antibodies | | | | | ELISA antibodies (IU) | |
|---|---|---|---|---|---|---|---|---|
| **Trial arm** | **N** | **CVB1 (titer)** | **CVB2 (titer)** | **CVB3 (titer)** | **CVB4 (titer)** | **CVB5 (titer)** | **Serion IgG** | **Serion IgM** |
| Placebo | 5 | 0<br>(0–1024) | 0<br>(0-1024) | 0<br>(0–2048) | 0<br>(0-0) | 0<br>(0-4096) | 3.82<br>(1.82-9.67) | 4.44<br>(4.06-8.47) |
| Low PRV-101 dose | 10 | 256<br>(64-8192) | 2<br>(0–3072) | 128<br>(6–8192) | 256<br>(32–4096) | 40<br>(0-2048) | 7.94<br>(1.32-19.34) | 6.29<br>(2.98-39.84) |
| High PRV-101 dose | 11 | 768<br>(128-2048) | 48<br>(0–6144) | 512<br>(128-8192) | 2048<br>(128-8192) | 1024<br>(12-4096) | 14.00<br>(2.46–24.14) | 5.48<br>(2.85-29.14) |

**Supplementary Table 2**. Median (range) neutralizing antibody titers (linear scale) against CVB1-5 viruses at the PROVENT-IIS time-point among study participants who were seronegative against the tested CVB type at baseline. The data has not been adjusted to the EOS time point.

| | | Neutralizing antibodies | | | | |
|---|---|---|---|---|---|---|
| **Trial arm** | **Participants (N for each CVB 1-5)** | **CVB1 (titer)** | **CVB2 (titer)** | **CVB3 (titer)** | **CVB4 (titer)** | **CVB5 (titer)** |
| Placebo | 4/3/3/5/3 | 0<br>(0 - 10) | 0<br>(0 – 0) | 0<br>(0 – 0) | 0<br>(0 – 0) | 0<br>(0 – 0) |
| Low PRV-101 dose | 9/8/6/9/7 | 256<br>(64 – 1536) | 1<br>(0 – 12) | 96<br>(6 – 128) | 256<br>(32 – 512) | 12<br>(0 – 192) |
| High PRV-101 dose | 10/10/9/7/3 | 896<br>(128 – 2048) | 40<br>(2 – 512) | 512<br>(128 – 2048) | 512<br>(128 – 4096) | 32<br>(12 – 192) |

| | Baseline (week 0) | | | | | IIS time point (week 101-120) | | | | |
|---|---|---|---|---|---|---|---|---|---|---|
| ARM | CVB1 | CVB2 | CVB3 | CVB4 | CVB5 | CVB1 | CVB2 | CVB3 | CVB4 | CVB5 |
| High | 0 | 0 | 0 | 0 | 0 | 192 | 18 | 128 | 256 | 12 |
| | 0 | 0 | 0 | 768 | 512 | 256 | 21 | 128 | 4096 | 2048 |
| | 8 | 0 | 256 | 0 | 2048 | 768 | 12 | 8192 | 512 | 4096 |
| | 0 | 0 | 0 | 0 | 256 | 1536 | 0 | 512 | 4096 | 3072 |
| | 0 | 0 | 0 | 1024 | 512 | 768 | 170 | 512 | 2048 | 4096 |
| | 0 | 0 | 0 | 512 | 0 | 2048 | 72 | 512 | 4096 | 192 |
| | 0 | 0 | 0 | 0 | 96 | 256 | 32 | 512 | 1024 | 1024 |
| | 0 | 1024 | 2048 | 0 | 32 | 3072 | 8192 | 4096 | 128 | 1024 |
| | 0 | 0 | 0 | 0 | 0 | 1024 | 256 | 384 | 512 | 32 |
| | 0 | 0 | 0 | 2048 | 4 | 576 | 0 | 512 | 8192 | 384 |
| | 0 | 0 | 0 | 0 | 128 | 2048 | 768 | 2048 | 4096 | 1536 |
| Low | 0 | 0 | 0 | 0 | 0 | 512 | 16 | 128 | 256 | 12 |
| | 0 | 0 | 0 | 0 | 0 | 72 | 0 | 6 | 32 | 16 |
| | 0 | 0 | 0 | 0 | 0 | 96 | 0 | 12 | 128 | 0 |
| | 1536 | 512 | 0 | 0 | 0 | 5461 | 3072 | 64 | 512 | 192 |
| | 0 | 0 | 1536 | 0 | 0 | 42 | 4 | 2048 | 256 | 6 |
| | 0 | 0 | 768 | 0 | 6 | 1152 | 4 | 8192 | 384 | 64 |
| | 0 | 0 | 0 | 2048 | 512 | 192 | 6 | 128 | 4096 | 2048 |
| | 0 | 1536 | 3072 | 0 | 0 | 384 | 2048 | 8192 | 192 | 8 |
| | 0 | 0 | 256 | 0 | 0 | 192 | 0 | 8192 | 512 | 128 |
| | 0 | 0 | 0 | 0 | 1536 | 256 | 0 | 128 | 256 | 2048 |

**Supplementary Figure 4.** Heatmap showing neutralizing antibody titers at baseline (left panel) and at IIS time point (right panel) per participant in the high and low dose treatment groups. Twofold serum dilutions were used to assess the level of neutralizing antibody titers against the tested viruses, 0 value indicating no neutralizing antibodies. Each row represents a study participant. The titers are presented as an average of two repeats. The adjusted IIS-titers come from the formula presented in statistical analyses section.

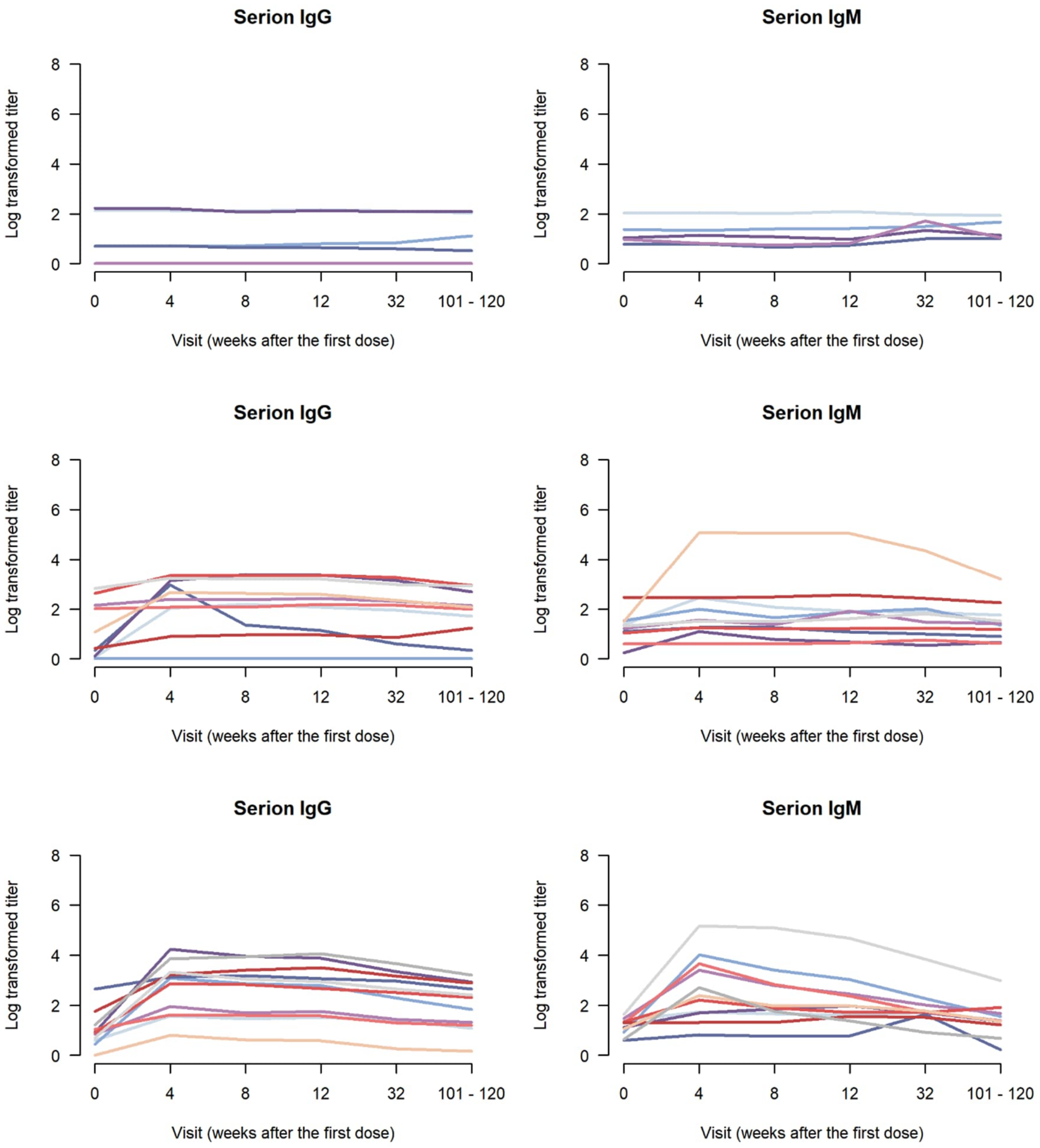


**Supplementary Figure 5**. The levels of IgG and IgM antibodies in ELISA assays in individual PROVENT-IIS trial participants. The top panel represents the placebo group, the middle panel the low dose group and the bottom panel the high dose group. Each participant is presented as an individual line. IIS values have been adjusted to those of the EOS time point.

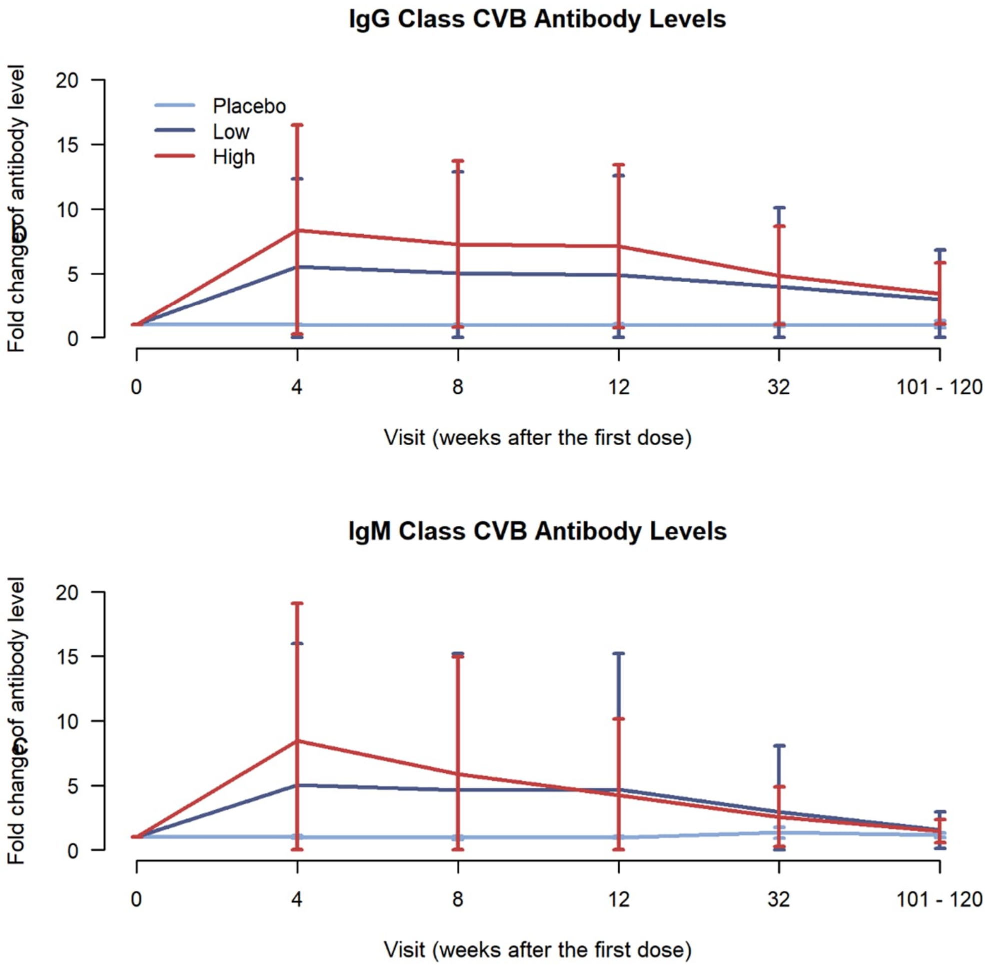


**Supplementary Figure 6**. Fold change (mean +/-SD) in IgG and IgM class CVB antibodies as compared to baseline values. IIS values have been adjusted to those of the EOS time point.
